# One size does not fit all: Single-subject analyses reveal substantial individual variation in electroencephalography (EEG) characteristics of antidepressant treatment response

**DOI:** 10.1101/2020.11.09.20227280

**Authors:** Gwen van der Wijk, Yaruuna Enkhbold, Kelsey Cnudde, Matt W. Szostakiwskyj, Pierre Blier, Verner Knott, Natalia Jaworska, Andrea B. Protzner

## Abstract

Electroencephalography (EEG) characteristics associated with treatment response show potential for informing treatment choices for major depressive disorder, but to date, no robust markers have been identified. Variable findings might be due to the use of group analyses on a relatively heterogeneous population, which neglect individual variation. However, the correspondence between group level findings and individual brain characteristics has not been extensively investigated. Using single-subject analyses, we explored the extent to which group-based EEG connectivity and complexity characteristics associated with treatment response could be identified in individual patients. Resting-state EEG data and Montgomery-Åsberg Depression Rating Scale symptom scores were collected from 43 patients with depression (23 females) before, at 1 and 12 weeks of treatment with escitalopram, bupropion or both. The multivariate statistical technique partial least squares was used to: 1) identify differences in EEG connectivity (weighted phase lag index) and complexity (multiscale entropy) between responders and non-responders to treatment (≥50% and <50% reduction in symptoms, respectively, by week 12), and 2) determine whether group patterns could be identified in individual patients. The group analyses distinguished groups. Responders showed decreased alpha and increased beta connectivity and early, widespread decreases in coarse scale entropy over treatment. Non-responders showed an opposite connectivity pattern, and later, spatially confined decreases in coarse scale entropy. These EEG characteristics were identified in ∼40-60% of individual patients. Substantial individual variation highlighted by the single-subject analyses might explain why robust EEG markers of antidepressant treatment response have not been identified. As up to 60% of patients in our sample was not well represented by the group results, individual variation needs to be considered when investigating clinically useful characteristics of antidepressant treatment response.

**Author summary:** Major depression affects over 300 million people worldwide, placing great personal and financial burden on individuals and society. Although multiple forms of treatment exist, we are not able to predict which treatment will work for which patients, so finding the right treatment can take months to years. Neuroimaging biomarker research aims to find characteristics of brain function that can predict treatment outcomes, allowing us to identify the most effective treatment for each patient faster. While promising findings have been reported, most studies look at group-average differences at intake between patients who do and do not recover with treatment. We do not yet know if such group-level characteristics can be identified in individual patients, however, and therefore if they can indeed be used to personalize treatment. In our study, we conducted individual patient analyses, and compared the individual patterns identified to group-average brain characteristics. We found that only ∼40-60% of individual patients showed the same brain characteristics as their group-average. These results indicate that commonly conducted group-average studies miss potentially important individual variation in the brain characteristics associated with antidepressant treatment outcome. This variation should be considered in future research so that individualized prediction of treatment outcomes can become a reality.

**Trial registration:** clinicaltrials.gov; https://clinicaltrials.gov; NCT00519428

## Introduction

Neuroscience research on major depressive disorder (MDD) has greatly improved our understanding of the brain alterations associated with MDD. An accumulation of studies comparing patients with MDD to healthy controls have highlighted both local and global alterations in brain network function, indicating that it may best be characterized as a network disorder [1, 2]. Studies have also examined relationships between brain network characteristics and antidepressant treatment success; his is especially relevant given the variability in treatment outcomes in MDD (e.g. [3]). Despite high hopes for the application of such research in clinical practice, findings have been variable and no robust diagnostic or prognostic information for individual patients have been reported to date [4, 5].

Functional connectivity, which is a commonly used measure of brain network function that measures the level of synchronized activity between brain regions, has shown promise for revealing network characteristics associated with treatment success [6-9]. Two electroencephalography (EEG) studies investigating associations between treatment success following 8 weeks of pharmacotherapy and functional connectivity found that weaker low frequency (delta, theta and alpha) connectivity at baseline, and a decrease in connectivity at these frequencies in right frontal and temporal electrode pairs was associated with better outcomes [6, 7]. However, increased alpha connectivity with treatment has also been associated with better treatment outcomes [9]. In the beta frequency band, some studies found lower pre-treatment connectivity and an early increase in connectivity, again mostly at frontal, temporal and central sites, to be associated with a better response [8, 9], while others did not find any treatment-related effects with beta [6], further highlighting the variability of connectivity findings in this context.

Researchers have also investigated network dynamics in MDD by examining complexity in brain signals, which provides complementary information to more traditional measures of brain network function [10]. Signals are considered to be complex when they have both stochastic and deterministic properties, and thus are neither completely predictable nor entirely random [11]. Some studies suggest that patients with MDD exhibit greater signal complexity than controls [12-15], and decreases in complexity have been associated with symptom improvement [16, 17]. In contrast, Čukić and colleagues [18] found higher complexity in patients in remission from MDD compared to both currently depressed patients and healthy controls. Importantly, most of these studies assessed complexity only at high temporal resolutions (1-10ms between datapoints). Our group found no association between treatment response and complexity at these fine temporal scales prior to treatment, but demonstrated that greater treatment response was associated with greater complexity at lower temporal resolutions [19].

These variable findings might be explained by the high degree of heterogeneity in patients with MDD [3, 20, 21]. Specifically, the studies reviewed above generally used group analyses to study small to moderate samples of patients with MDD, which could easily lead to variable findings if extensive individual variation exists in brain data recorded from this population. Group analyses tend to capture central tendencies in the data and treat individual variation outside of these common features as noise, and might therefore highlight different commonalities depending on the sample of patients included in each study. Evidence of such individual divergence from group level findings in brain recordings was recently shown within a relatively homogeneous sample of healthy young adults, where individual participants had qualitatively different brain network organization compared to the group-average estimate [22]. It is likely that similar (or even greater) individual variation in brain network characteristics exist in patients with MDD and could be a factor in the variable findings discussed above, but this has yet to be investigated. Defining the extent to which group findings are representative of brain features at an individual level in the context of MDD could inform future attempts to apply group findings to individuals.

Here we explored the extent to which group level findings were representative of individuals within the same sample by examining EEG connectivity and complexity. Similar to other studies in this field (e.g. [8, 9, 18]), our sample consisted of a moderate sample of 43 patients, receiving multiple antidepressant medication regimens (in our case, escitalopram, bupropion or their combination) for 12 weeks. For group analyses, patients were divided into responders (≥50% symptom improvement on the Montgomery-Åsberg Depression Rating Scale [MADRS] from baseline to 12 weeks of treatment) and non-responders (<50% improvement). EEG was measured at baseline, and after 1 and 12 weeks of treatment. We identified patterns of change in EEG connectivity and complexity at the group level in responders and non-responder, and examined the extent to which each individual’s results conformed to their own group’s pattern (e.g., responders showing the responder pattern) through single-subject analyses. Based on the most consistently reported findings from previous literature, we expected that responders would exhibit decreased connectivity at lower frequencies with treatment, and decreased complexity in response to pharmacotherapy [6-8, 17]. We expected that individual responders would generally follow the pattern found in the responder group, while non-responders might show more variable changes.

## Methods

### Participants

Fifty-three adults with a primary diagnosis of major depressive disorder (MDD), as assessed by a psychiatrist with the Structured Clinical Interview for DSM-IV-TR [SCID-IV-TR] [23], participated in this study, as previously described [19]. Briefly, patients were excluded if they had any other Axis I disorder (except for anxiety disorders), recent (< 6 months ago) problems with substance abuse/dependence, an unstable medical condition, significant suicide risk, seizure history, or if they had been previously treated for their current depressive episode with the current study medications. Medicated patients underwent a supervised washout period prior to study commencement (>5 weeks for fluoxetine, 1 week for other medications). As part of a larger clinical trial conducted between August 2007 and March 2012 [24], patients received either escitalopram (ESC) and placebo, bupropion (BUP) and placebo, or a combination of the two medications for 12 weeks. Assignment to a specific treatment regimen was randomized (double blind).

Depressive symptoms were assessed using the Montgomery-Åsberg depression rating scale [MADRS] [25, 26], every week during the first 4 weeks, and biweekly for the remaining 8 weeks. Dosage was increased if tolerated, and remission was not yet reached (average dose at 12 weeks for the current sample: dual treatment: ESC = 32 mg, BUP = 379 mg; monotherapy: ESC = 34 mg, BUP = 425 mg). All patients had a baseline MADRS score ≥ 22. Patients whose MADRS scores improved ≥50% from baseline to 12 weeks were considered responders (R), while those who improved <50% were considered non-responders (NR). Due to participant drop-out and issues with EEG data quality, 10 participants were excluded from the current study, leaving 43 participants for data analysis (i.e. complete datasets at baseline, week 1 and 12), of whom 25 were responders and 18 were non-responders. Demographic and clinical characteristics can be found in **Table 1**. Statistical tests were performed in Excel to check for differences between responder and non-responder groups. All participants provided written informed consent, and were reimbursed $30 CAD/testing session. This study was approved by the Royal Ottawa Health Care Group and University of Ottawa Social Sciences & Humanities Research Ethics Boards.

**Table 1.** Demographic and Clinical Characteristics (Means ± Standard Error) of Antidepressant Treatment Responders and Non-responders

|  | **Responders**<br>**(N = 25)** | **Non-responders**<br>**(N = 18)** | **Statistics** |
| --- | --- | --- | --- |
| *Sex (F/M)* | 14/11 | 9/9 | $\chi^2(3) = 0.15, p = .70$ |
| *Age* | 35.1 ± 2.1<br>(range: 19-57) | 44.8 ± 2.7<br>(range: 20-63) | $t(35) = 2.77, p = .009^*$ |
| *Education (years)* | 15.4 ± 0.5 | 16.3 ± 0.6 | $t(34) = 1.20, p = .24$ |
| *Ethnicity* | 22 Caucasian; 2 Asian; 1 South Asian | 17 Caucasian; 1 African | $p = .48$ (Fisher's exact test) |
| *Comorbid anxiety (Yes/No)* | 3/22 | 3/15 | $p = .68$ (Fisher's exact test) |
| *Treatment regimen*<br>*(ESC+BUP/BUP+placebo/*<br>*ESC+placebo)* | 12/6/7 | 5/6/7 | $\chi^2(5) = 1.79, p = .41$ |
| *Baseline MADRS score* | 29.4 ± 0.9 | 32.2 ± 1.1 | $t(36) = 1.91, p = .064$ |
| *MADRS score at 1 week* | 23.1 ± 1.6 | 27.9 ± 1.9 | $t(37) = 1.92, p = .063$ |
| *MADRS score at 12 weeks* | 6.0 ± 1.0 | 24.9 ± 1.9 | $t(26) = 8.85, p < .001^*$ |

Group differences were examined using independent samples t-tests in Excel, unless reported otherwise. \*
Significance at $p < .05$ . F = female, M = male, ESC = escitalopram, BUP = bupropion, MADRS = Montgomery-Åsberg Depression Rating Scale

### EEG data collection

Resting state EEG recordings were collected before the start of treatment (baseline), 1 week and 12 weeks after treatment initiation. Participants abstained from caffeine and nicotine >3 hours prior to testing, and did not take any drugs, other than the prescribed antidepressants, on the nights before testing, except if needed for a stabilized medical condition. Two 3-minute resting-state EEG recordings were collected, one with eyes open (EO) and one with eyes closed (EC), while participants sat in a temperature- and light-controlled testing chamber. Ip et al. (2018) showed that 3-minute EEG recordings are enough to extract reliable EEG characteristics in the theta, alpha and beta bands using a test-retest design. The order of EO and EC testing was counterbalanced between participants and sessions. EEG was recorded using 32 Ag/AgCl electrodes embedded in a cap (EasyCap, Inning am Ammersee, Germany), with electrodes positioned according to a variant of the 10-20 system [27]. AFz served as the ground, and the average of the two mastoid channels (Tp9/Tp10) was used as the reference. Four additional channels were placed outside the left and right eye canthi, and above and below one eye, to monitor electrooculographic (EOG) activity. Data was sampled at 500 Hz, and impedance was <5KΩ (BrainVision Recorder, Gilching, Germany).

### EEG Preprocessing

EEG data were preprocessed using EEGLAB v13.4.4b [28] in MATLAB 2014 (The MathWorks, Inc., Natick, Massachusetts). Raw EEG data were bandpass filtered (0.5-55 Hz; slope: 12 dB/octave) using ERPlab’s IIR butterworth filter, notch filtered at 60 Hz (lower and upper edge: 55-65 Hz) using EEGlab’s basic FIR filter and segmented into 2s epochs. We used an independent component analysis (ICA) to identify and eliminate noise and ocular artifacts. Channels with excessive noise or drift were excluded from the ICA procedure, and subsequently interpolated using EEGlab’s spherical spline interpolation function. No more than two channels were interpolated for each participant and session. Epochs were visually inspected following ICA, and those with remaining artifacts were manually rejected. An average of 85.7 (range: 63-113) artifact free epochs were obtained per participant, state (EC/EO) and session (baseline, week 1 & 12), including 28 electrodes (Fp1/2; F3/ 4; F7/8; FC1/2; FC5/6; C3/4; CP1/2; CP5/6; P3/4; P7/8; T7/8; O1/2; Fz/Cz/Pz/Oz). The two reference channels, and two additional channels that were consistently flat for each participant (FT9/FT10), were excluded from analysis. There was no statistical difference between responders and non-responders in number of artifact-free epochs per session or state (*p-*values: .09-.9).

### Connectivity analysis

Functional connectivity, as quantified by the weighted phase lag index (WPLI), was calculated for each unique combination of the 28 channels (378 pairs) using the open source Fieldtrip toolbox [29] in MATLAB. WPLI is a modified version of the phase lag index (PLI), which was first described in 2007 by Stam and colleagues [30]. It estimates connectivity by calculating the phase angle difference between EEG signals from two channels for each time point, and determining the consistency in these phase lags over time. As such, if the difference in phase between two channels is similar over time, the PLI will be high, indicating high connectivity between two channels. An advantage of the PLI compared to other EEG connectivity measures is that it is less sensitive to volume conduction, because it disregards any phase lags of 0 and π. The WPLI also takes into account that phase lags can easily turn into leads and vice versa (e.g. a slightly positive phase angle difference can turn into a slightly negative phase angle difference). While the PLI is sensitive to such small disturbances in phase lags, the WPLI resolves this issue by giving greater weight to angle differences around 0.5π and 1.5π [31]. The result is a value between 0 and 1, with higher values indicating stronger connectivity. WPLI was calculated in two ways, first across epochs, as is commonly used and enables direct comparisions with previous findings, and then within epochs, which is less commonly used, but enables single-subject analyses.

#### Across-epoch WPLI

Phase information was first extracted for each epoch, channel and frequency bin (0.5-50 Hz, 0.5 Hz bins) using Fieldtrip’s fast Fourier transformation algorithm. A Hanning taper was used for the lower frequencies (0.5-30 Hz), while a multi-taper using the dpss (discrete prolate spheroidal sequences) method with 2 Hz smoothing was applied to the higher frequencies (31-50 Hz), to optimize sensitivity of spectral content at each frequency. WPLI values were then calculated by considering the consistency of phase lags over epochs at each frequency bin for all channel pairs using Fieldtrip’s connectivity function.

#### Single-epoch WPLI

This across-epoch method is not suitable for single-subject analyses, because it does not allow calculation of WPLI for individual epochs. Therefore, a second, less common approach was used to calculate WPLI for the single-subject analyses [32]. Instead of extracting one phase value per epoch, phase was determined for each time point within an epoch using a time-frequency transformation with Morlet wavelets in the time domain. To have reasonable temporal and frequency resolution, the length of the wavelets was increased with frequency in regular steps, from 3 cycles at 4 Hz to 7 cycles at 50 Hz [32]. Frequencies below 4 Hz were not included as the length of our epochs (2 seconds) was too short to provide reliable estimations at these frequencies (i.e. 3 cycles of a 1 Hz wavelet are longer than 2 seconds). WPLI could then be calculated for individual epochs by examining the consistency of phase lags over time points within each epoch [32]. These individual epoch data were used for the individual connectivity analyses. To confirm that this single-epoch approach provides similar results at the group level as the across-epoch approach, we also averaged these data over epochs for each participant, and ran the exact same group level analyses.

### Brain signal complexity analysis

Multiscale entropy (MSE) was used to quantify brain signal complexity. An advantage of MSE over other measures of complexity is the incorporation of multiple time scales. This feature is important because it differentiates between signals that are purely random (such as white noise) and those comprised of both random and deterministic components (such as 1/f or coloured noise). Signals that are purely random show a rapid decline in the MSE curve with increasing scale whereas those with temporal inter-dependencies will have a more gradual shift in the MSE curve [33, 34]. A detailed description and theoretic background for MSE is outlined in Costa and colleagues [11]. In short, MSE estimates the regularity of a signal by evaluating the ratio of similar patterns of different lengths repeating over several time scales. It is calculated in two steps. First, the raw signal is resampled several times to create data sequences that represent different temporal scales. Essentially, an increasing number of non-overlapping data points are averaged into one new data point. The first timescale is the (cleaned) raw time series. With a sample rate of 500Hz in the current study, time scale 1 had a temporal resolution of 2 milliseconds between data points. For time scale 2, two consecutive data points were averaged, yielding a temporal resolution of 4 milliseconds; for time scale 3, averaging occurs over three time points yielding a temporal resolution of 6 milliseconds, and so on. The coarsest scale used in this study was 20 (temporal resolution of 40 milliseconds).

Next, sample entropy is calculated at each time scale. Sample entropy determines the natural logarithm of the ratio of patterns of length *m* over patterns of length *m+1* repeated within one epoch. This gives a value between 0 and 1, with higher numbers indicating a less predictable/more variable signal (i.e. fewer patterns of length *m+1* compared to the number of patterns of length *m*). In line with previous studies, (e.g. [19, 35]) and guidelines outlined by Richman and Moorman [36], parameter *m* was set to 2 in this study, while the similarity criterion *r*, which determines which points in the time series are considered to be ‘the same’, was set to 0.5 (i.e. two data points were treated as indistinguishable if their amplitudes differed <50% of the standard deviation of the time series). MSE was calculated for each epoch and electrode at each time scale, using the algorithm available at www.physionet.org/physiotools/mse/. Single epoch MSE data were used for statistical analyses at the individual level. MSE values were also averaged over epochs to provide one MSE value for each electrode and time scale per participant, session (baseline, week 1 and 12) and state (EC and EO), which were used for group level analyses.

### Regression of age effects

To control for differences in age between responder and non-responder groups (see **Table 1**), and because both brain signal complexity and connectivity have been observed to change with age [19, 37-40], age was regressed out of the data before the statistical group comparisons using an in-house MATLAB script [41, 42].

### State contrasts

Consistent with MSE and connectivity differences between EO and EC states observed in previous studies [19, 43-46], we found strong EO/EC contrasts in our analyses that masked changes occurring over assessment sessions (see **Figure S1 & S2**). Therefore, we performed analyses on EO and EC data separately. To maximize the chance of replicating group findings at the individual level, we performed single-subject analyses on the data showing the strongest effects (EC for WPLI, EO for MSE), and present those group findings below (other group findings are presented in **Supplementary Materials** [**Figure S3 & S4]**).

### Statistical analyses with PLS-SVD

Partial least squares with singular value decomposition (PLS-SVD) is a multivariate statistical approach that can detect condition- and/or group-related differences in whole-brain variables [47, 48]. Briefly, PLS-SVD calculates the between-subject covariance between experimental design characteristics (in this case, responder status and assessment sessions) and brain characteristics (in this case, WPLI or MSE). Then, this covariance matrix is decomposed using SVD into orthogonal latent variables (LVs) that account for most of the covariance between groups/conditions and brain characteristics, revealing the optimal associations between specific groups/conditions and spatiotemporal patterns in the brain. LVs contain several components. One is the singular value, which indicates the strength of the effect the LV represents. Another component holds the element loadings, which represent the pattern of the specific data elements (in this study frequencies and electrode pairs for WPLI, and time scales and electrodes for MSE) that show the given contrast. These element loadings are used to compute brain scores: the dot product of the element loadings with each participants’ data for each assessment session. Brain scores represent the extent to which each participant expresses the given contrast in a single number per participant, and can therefore be used to get an overview of the contrast between groups/conditions.

Statistical testing occurs at two levels in PLS-SVD analyses. First, the overall significance of the LV is determined using permutation tests. In each permutation, the data are randomly shuffled between conditions (within participants) and between groups, and PLS-SVD analysis is performed on the shuffled data just as on the actual data. LVs are considered significant when their singular value is more extreme than 95% of the singular values calculated from the randomly shuffled data (corresponding to p < .05). In the current study, 500 permutations were performed for each analysis. Second, the stability of the identified pattern over participants is established through bootstrap resampling. In essence, the PLS-SVD analysis is repeated with different subsamples of participants, to see how consistently each electrode pair/electrode and frequency/time scale display the identified pattern of differences across the whole sample. This consistency is quantified as a bootstrap ratio, which is calculated by dividing the element loadings by the standard error of the created bootstrap distribution for each element. In addition to determining the stability of the pattern, bootstrap resampling also protects against the influence of outliers, as subsamples with and without the outlier would produce different outcomes, thereby decreasing the consistency of the findings (i.e. the bootstrap ratio). In practice, this means that effects that are driven largely by an outlier get attenuated. Bootstrap ratios are similar to z-scores, with absolute values ≥ 3.1 corresponding to ∼99% confidence interval. In this study, bootstrap resampling was performed 200 times. As each statistical test is computed in one mathematical step, no correction for multiple comparisons is necessary [47]. P-values indicating significance levels, and percentage of crossblock covariance explained (PCCE) are reported for each LV of interest. PLS-SVD analyses were applied both at a group and individual level.

### Group level analyses

Both groups (responders vs. non-responders) and all sessions (baseline, 1 & 12 weeks of treatment) were entered in four PLS-SVD analyses: two for connectivity (EO/EC states separately) and two for complexity (EO/EC). As all showed interaction effects between groups and assessment sessions, two additional analyses were run for each analysis for responders and non-responders separately, again including all sessions. The p-values of these follow-up analyses were corrected for multiple comparisons using the Bonferroni method.

The input data consisted of across-epoch WPLI/averaged MSE values, organized into 2D matrices with n ^*^ k rows, and m ^*^ t columns, with n being the number of participants (R: 25; NR:18) and k the number of conditions (assessment sessions: 3). M and t represent the spatiotemporal elements, namely the number of electrode pairs (378) and frequencies (99) for the WPLI analyses and the number of electrodes (28) and timescales (20) for the MSE analyses. The same procedure was followed for the averaged, single-epoch WPLI data (*Methods – Connectivity analyses*).

### Single-subject analyses

Non-rotated (hypothesis-driven) PLS-SVD analyses were performed for each individual, using single-epoch EC WPLI data, and single-epoch EO MSE data. Single epoch WPLI/MSE values were organized into similar 2D matrices as described for the group analyses, only now each participant had their own datamat, with the n dimension representing the number of epochs instead of participants [48]. Non-rotated PLS-SVD was chosen because it allows one to determine whether and to what extent a specific, predefined contrast is present in the data. Using the contrasts found in the group analyses, non-rotated PLS-SVD was used to test whether each individual followed the pattern of change observed in responder and non-responder groups. No correction for multiple comparisons was applied, as these analyses aimed to replicate group findings in separate datasets for each individual.

The similarity of the individual PLS-SVD outcomes to the group PLS-SVD outcomes was quantified in two ways. First, the similarity was estimated quantitatively, by correlating the stable (|BSR| > 2, corresponding to ∼95% confidence interval) element loadings (i.e. the spatiotemporal brain pattern) of the group results with the element loadings of each participants’ individual analysis in MATLAB. For connectivity, the element loadings from the group analyses on averaged single-epoch WPLI were used for this correlation procedure (*Methods – Connectivity analyses)*. If participants did not significantly show the predefined contrast (responder/non-responder), indicating the timing and direction of the change highlighted by element loadings, their results were not correlated with the results of that group and were included as ‘showing no correlation with the group pattern’ in the summaries. Second, to balance arbitrary cut-offs and mimic clinical interpretability, significant individual outcomes were visualized and classified by two independent raters, blind to response status, as being similar to either or both the responder or non-responder group patterns, or neither. Important responder and non-responder features were selected based on visual inspection of the most consistent changes across time (i.e. those with |BSR| > 3.1) in the group analyses. The percentage of participants showing moderate-strong correlations (r≥.4; [49]) with their own group outcome and/or being classified as conforming to their own group pattern exclusively was determined as an indicator of the replicability of the group patterns at the individual level.

## Results

### Participants

By design, responders had lower MADRS scores at week 12, but not at baseline or week 1 (**Table 1**). Apart from responders being younger than non-responders, the two groups did not differ statistically in clinical and demographic characteristics (**Table 1**). We accounted for the age difference by regressing age effects out of our data before running the statistical tests at the group level.

### Group analyses - WPLI

The PLS-SVD analysis including both groups and all sessions identified one significant LV (*p* < .001, PCCE = 35.07%). As this LV presented an interaction effect between groups and sessions, two additional analyses for each group separately were run, with the statistical significance threshold corrected to α < .025. These analyses revealed a complex, opposite pattern of change from weeks 1 to 12 in responders (*p*=.024, PVE=55.8%) and non-responders (*p* = .032, PCCE=56.6%; **Figure 1**), although the non-responder LV only approached significance. The most prominent frequencies for each group are highlighted by red boxes in **Figure 1**: Non-responders showed a widespread increase in alpha connectivity (10Hz), while responders exhibited an extensive increase in beta connectivity (22Hz). Considering the same frequencies in the opposite groups (e.g. alpha in responders; highlighted by blue boxes) revealed more spatially contained changes in the opposite direction: Responders showed a decrease in connectivity at 10Hz, while non-responders showed a decrease at 22Hz. In both groups, changes in alpha connectivity were most pronounced at interhemispheric frontal-to-occipito-parietal electrode pairs but involved additional electrode pairs in non-responders. The most consistent beta changes occurred in left intra-hemispheric connections in both groups, but also included right central and parietal electrode pairs in responders (**Figure 2**).

**Figure 1.**
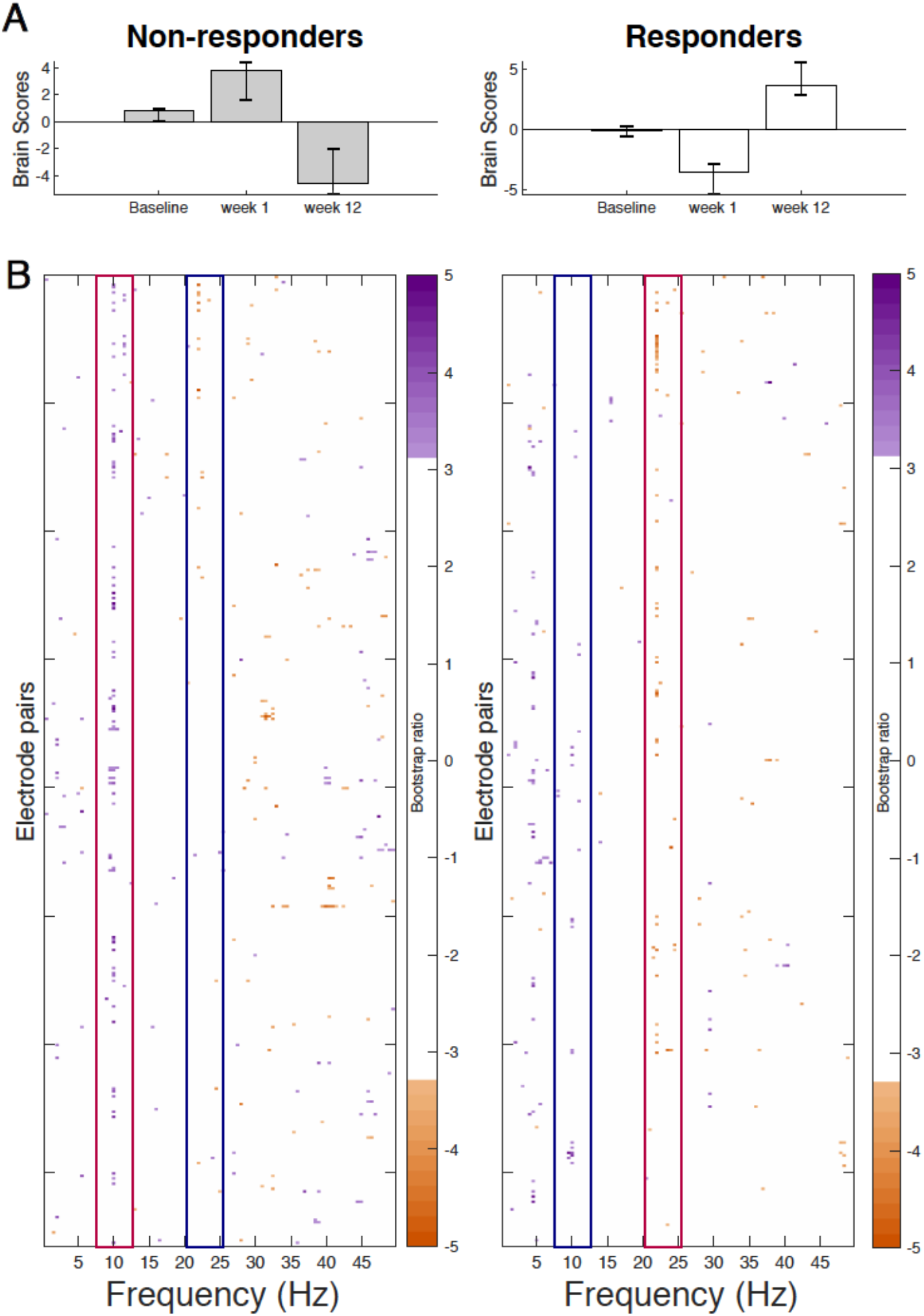
Results from the task partial-least squares (PLS-SVD) analyses examining change in connectivity as measured by weighted phase lag index (WPLI) over the course of antidepressant medication treatment in eventual non-responders (left) and responders (right). Bar graphs (A) depict the contrast between assessment sessions within groups, that was significantly expressed across each data set as determined by permutation testing. The statistical image plots (B) present the bootstrap ratio maps over all electrode pairs (rows) and frequencies (columns). The orange and purple pixels display where the contrast represented by the bar graphs was most reliable across participants as determined by bootstrapping. Positive values (purple) indicate increased WPLI in responders, and decreased WPLI in non-responders from 1 to 12 weeks of treatment, while negative values (orange) indicate decreased WPLI in responders and increased WPLI in non-responders from weeks 1 to 12. To aid interpretability, the most prominent increases in WPLI are highlighted by red boxes, while decreases in WPLI are outlined by blue boxes. As highlighted by these boxes, non-responders showed an increase in alpha and a decrease in beta connectivity from week 1 to week 12 of treatment, while responders showed the opposite pattern.

**Figure 2.**
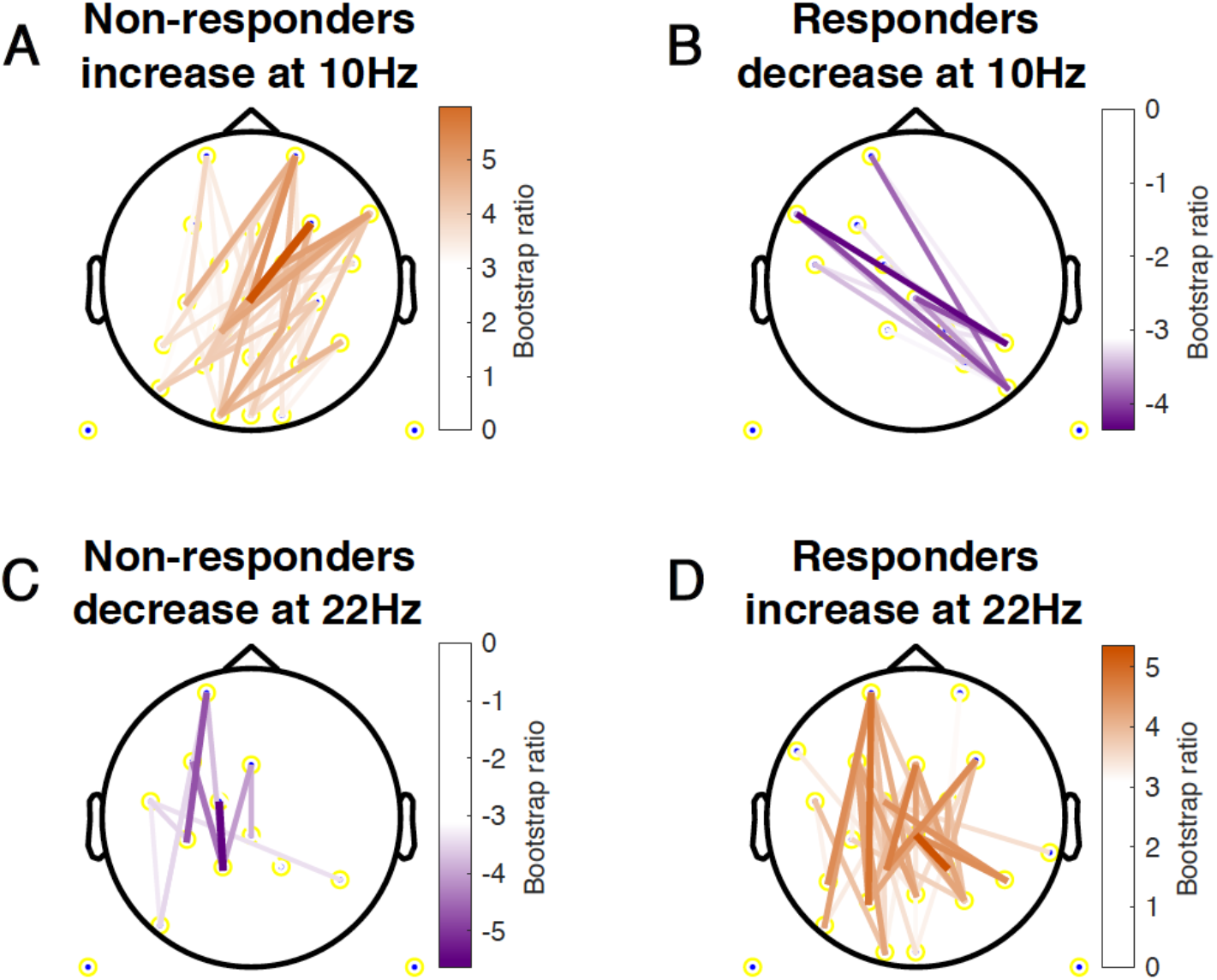
Topographical location of electrode pairs showing the most consistent change in connectivity as measured by weighted phase lag index (WPLI) over assessment sessions in non-responders (A/C) and responders (B/D) at 10Hz (A/B) and 22Hz (C/D). Positive values (orange) indicate increased WPLI from 1 to 12 weeks of treatment, while negative values (purple) indicate decreased WPLI from weeks 1 to 12.

The group PLS-SVD analyses performed on the averaged single-epoch WPLI data showed a similar pattern of change in connectivity in responder and non-responders (**Supplementary Material & Figure S5**). However, the results spread over multiple frequencies (e.g. from 8-14 Hz instead of dominantly at 10 Hz), which is unsurprising, considering the reduced spectral resolution associated with sliding window approaches [32].

### Group analyses - MSE

The PLS-SVD analysis examining changes in MSE over time in responders and non-responders identified one significant LV (*p* < .001, PCCE = 82.71%), which revealed an interaction effect. The analyses exploring changes for each group separately each found one significant LV (responders: *p* = .006, PCCE = 93.85%, non-responders: *p* = .02, PVE = 86.9%, significant at α < .025). Both groups showed a decrease in coarse scale complexity from baseline to 12 weeks, but the timing and extent of change differed. Responders showed an early (starting at week 1) and widespread decrease in coarse scale complexity, while non-responders showed a later (only present at week 12) decrease in coarse scale complexity in limited electrodes (**Figure 3** & **4**). Additionally, fine scale complexity increased only in non-responders.

**Figure 3.**
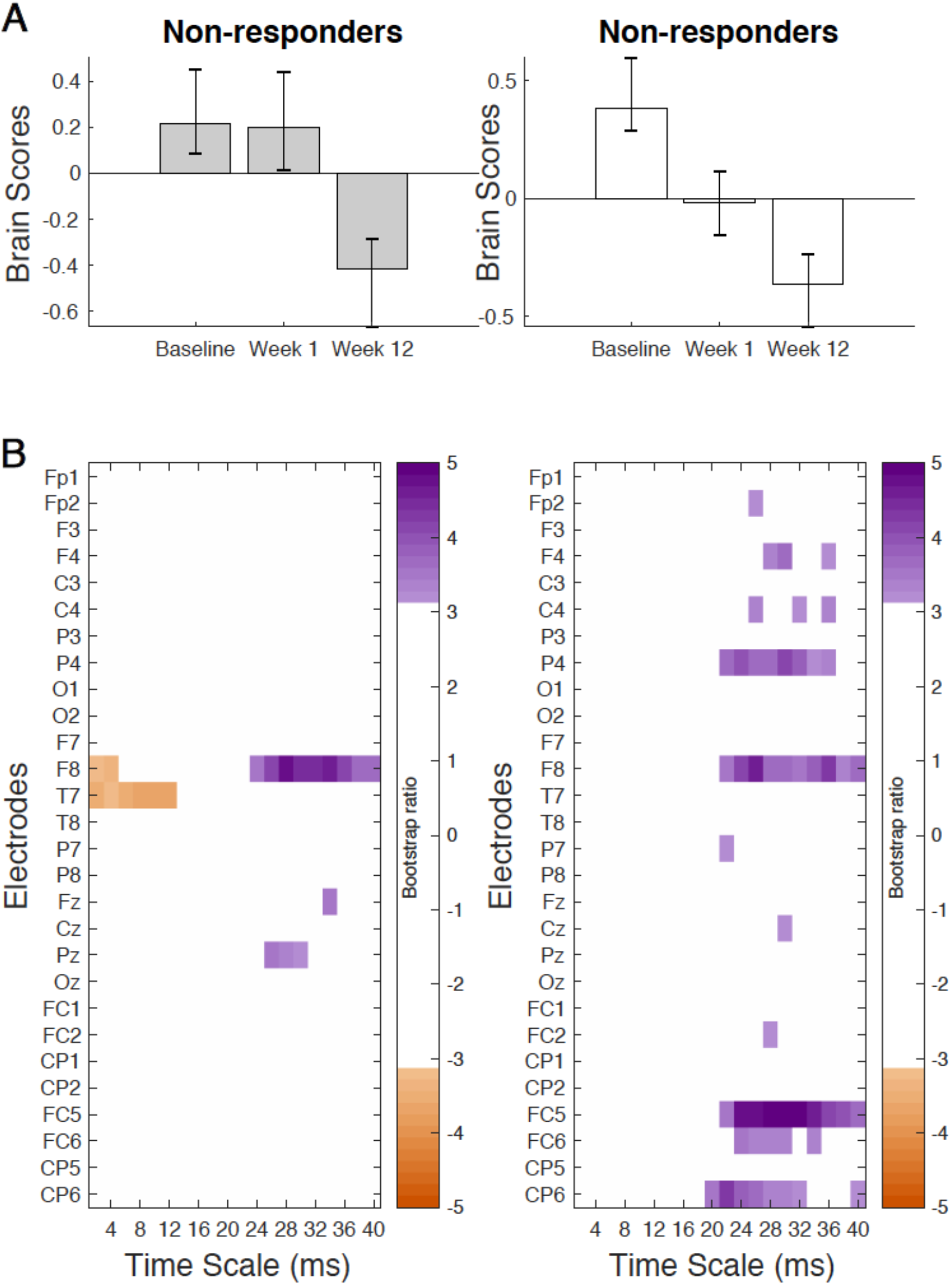
Results from the task partial-least squares (PLS-SVD) analyses examining change in complexity as measured by multiscale entropy (MSE) over the course of antidepressant medication treatment in non-responders (left) and responders (right). Bar graphs (A) depict the contrast between assessment sessions within groups, that was significantly expressed across each data set as determined by permutation testing. The statistical image plots (B) present bootstrap ratio maps over all electrodes (rows) and time scales (columns). The colored values display where the contrast represented by the bar graphs was most consistent across participants as determined by bootstrapping. Positive values (purple) indicate decreased MSE, while negative values (orange) indicate increased MSE at week 12 compared to baseline and week 1 in non-responders, and at week 12 compared to baseline in responders.

**Figure 4.**
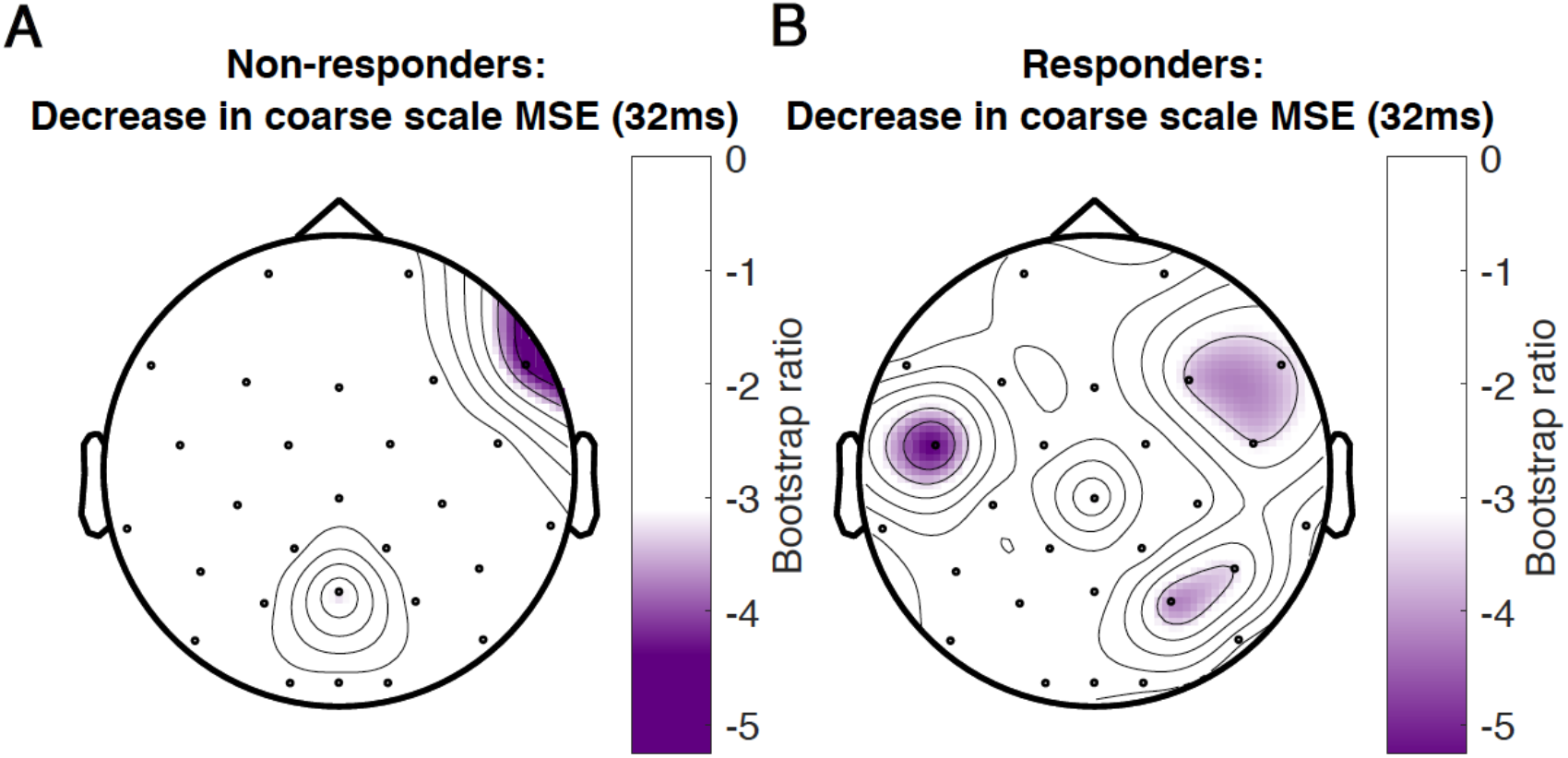
Topographical location of electrodes showing the most consistent change in complexity as measured by multiscale entropy (MSE) over assessment sessions in non-responders (A) and responders (B) at a time scale of 32ms between data points. Negative values (purple) indicate decreased MSE at week 12 compared to baseline and week 1 in non-responders, and decreased MSE at week 12 compared with baseline in responders. There was no increase in MSE for any electrode at this time scale.

### Individual analyses - WPLI

The pattern of change across assessments identified by the group PLS-SVD examining connectivity was similar regardless of the approach used to calculate WPLI, namely, consisting of a change in connectivity from week 1 to week 12 in both non-responders and responders (**Figure 1** & **S5**). Therefore, we applied this pattern in the non-rotated single-subject PLS-SVD analyses. As the non-responder and responder patterns only differed in the direction of change (i.e. increase or decrease in WPLI), only one contrast was defined for each analysis (0 1 −1). This contrast examines changes in WPLI from week 1 to week 12 but leaves the direction of change and at which frequencies this occurs to be determined by the data. All non-responders and 22/25 responders exhibited the predefined pattern of change at an uncorrected significance level (all *p* < .05), of whom 33 (19R/14NR) survived Bonferroni correction (*p* < .001).

The correlation procedure showed that 60.5% of individual patients exhibited moderate-strong positive correlations (i.e. r≥.4; [49]) between their individual and group outcomes. Another 9.3% showed weak positive correlations (i.e. .1<r<.4), while 14.0% revealed negative correlations between individual and group PLS-SVD outcomes. The remaining 16.3% of patients’ individual analyses either correlated negligibly (-.1<r<.1) or did not reach significance and were therefore not correlated (**Figure 5**).

**Figure 5.**
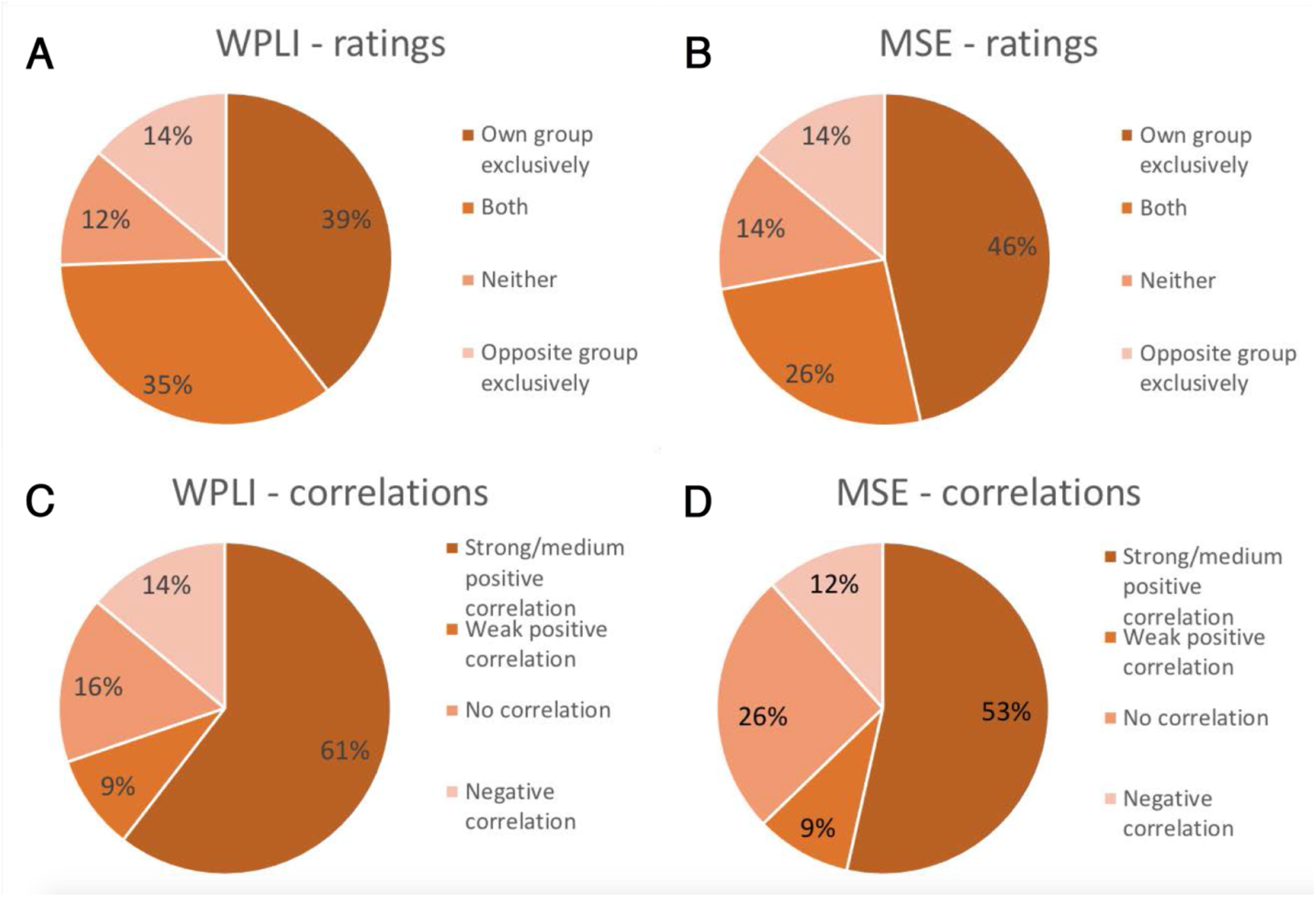
Proportion of participants that exhibited the same pattern as their own group exclusively, the pattern of the opposite group exclusively, both group patterns or neither group pattern in their individual analysis of connectivity as measured by weighted phase lag index (WPLI; A) and complexity as measured by multiscale entropy (MSE; B). Proportion of participants showing a strong/medium positive correlation (r≥.4), a weak positive correlation (.1<r<.4), a negligible or lack of correlation (-.1<r<.1) or a negative correlation (r<-.1) between their individual outcome matrix and that of their own group for WPLI (C) and MSE (D).

Following the direction of change found at 10Hz (alpha) and 22Hz (beta) for the responder and non-responder groups, individuals showing *a pattern of a meaningful decrease in alpha (∼8-14Hz) and/or increase in beta (∼18-30Hz) WPLI from week 1 to 12* were considered to fit the responder pattern, while patients showing *a pattern of a meaningful increase in alpha and/or decrease in beta WPLI from week 1 to 12* were considered to fit the non-responder pattern. Two authors examined each individual outcome pattern and independently decided whether they conformed to the outlined definitions. Inter-rater reliability was quantified using Cohen’s Kappa: ***κ***=0.76 for rating whether patients fit the responder pattern, and ***κ***=0.57 for rating whether patients fit the non-responder pattern. Raters discussed discrepancies until consensus, which indicated that 39.5% of patients fit the pattern of their own group exclusively, 34.9% fit both patterns, 14.0% only showed the pattern of the opposite groups, and the remaining 11.6% did not conform to either pattern (7% did not reach significance). **Table 2** shows the characteristics of patients who were rated as fitting their own WPLI pattern exclusively versus the other categories. Aside from individual responders conforming to the responder group pattern showing higher correlations with their own WPLI group pattern compared to responders assigned to other categories (p = .005, did not survive Holm’s sequential Bonferroni test for multiple comparisons), there were no obvious clinical and demographic differences between individuals in the different responder and non-responder categories.

**Table 2.** Demographic and Clinical Characteristics (Means ± Standard Error) of Antidepressant Treatment Responders and Non-responders Divided Based on which Individuals Matched the Group WPLI Patterns

|  | **Responders**<br>**(N = 25)** |  | **Non-responders**<br>**(N = 18)** |  |
| --- | --- | --- | --- | --- |
|  | *Responder pattern*<br>*only (N = 10)* | *Other categories*<br>*(N = 15)* | *Non-responder*<br>*pattern only (N = 7)* | *Other categories*<br>*(N = 11)* |
| *Sex (F/M)* | 4/6 | 10/5 | 4/3 | 5/6 |
| *Age* | 38.0 ± 3.7<br>(range: 23-57) | 32.5 ± 2.5<br>(range: 19-46) | 40.3 ± 4.6<br>(range: 20-57) | 47.6 ± 3.3<br>(range: 28-63) |
| *Education (years)* | 16.7 ± 0.8* | 14.5 ± 0.6 | 15.1 ± 0.8 | 17.1 ± 0.9 |
| *Ethnicity* | 9 Caucasian; 1<br>Asian | 13 Caucasian; 1<br>Asian; 1 South<br>Asian | 7 Caucasian | 10 Caucasian; 1<br>African |
| *Comorbid anxiety (Yes/No)* | 0/10 | 3/12 | 0/7 | 2/9 |
| *Treatment regimen*<br>*(ESC+BUP/BUP+placebo/*<br>*ESC+placebo)* | 4/2/4 | 8/4/3 | 2/2/3 | 3/4/4 |
| *Baseline MADRS score* | 29.4 ± 1.4 | 29.4 ± 1.3 | 33.1 ± 1.0 | 31.5 ± 1.7 |
| *MADRS score at 1 week* | 23.9 ± 2.8 | 22.6 ± 2.0 | 29.7 ± 2.6 | 26.7 ± 2.7 |
| *MADRS score at 12 weeks* | 5.1 ± 1.5 | 6.7 ± 1.3 | 25.4 ± 3.1 | 24.5 ± 2.5 |
| *Correlation with own group*<br>*WPLI pattern (>.4/<.4)* | 9/1* | 6/9 | 3/4 | 3/8 |
| *Correlation with own group*<br>*WPLI pattern* | 0.67 ± 0.09** | 0.13 ± 0.15 | 0.51 ± 0.07 | 0.30 ± 0.09 |
| *Correlation with own group*<br>*MSE pattern* | 0.44 ± 0.16 | 0.26 ± 0.12 | 0.40 ± 0.15 | 0.40 ± 0.18 |

Group differences were examined using independent samples t-tests and Fisher's exact test in Excel. \*\*
Significant differences between groups at $p < .05$ (with Holm's sequential Bonferroni test to correct for multiple comparisons, as these were unplanned comparisons). \* Group differences with $p < .05$ that did not survive the correction. F = female, M = male, ESC = escitalopram, BUP = bupropion, MADRS = Montgomery-Åsberg Depression Rating Scale

### Individual analyses - MSE

Non-rotated PLS-SVD was performed for all participants with two predefined contrasts: 1 0 −1 as the responder pattern (change from baseline to week 12, top of **Figure 3**) and 1 1 −2 as the non-responder pattern (no change from baseline to week 1, change from weeks 1 to 12, top of **Figure 3**). While all individual analyses revealed significant results for both predefined LVs (all *p* < .001), 16% of patients did not show the same pattern of change over assessment sessions as defined in their own group contrast.

The correlation procedure showed that 53.5% of individual outcome patterns correlated positively with moderate-high strength (r ≥ .4; [49]) to that of their groups. Another 9.3% showed a weak (.1 < r < .4) positive correlation between their individual outcome and that of their group. Of the remaining individuals’ analyses, 11.6% yielded negative correlations, while 25.6% either showed negligible correlations (-.1 < r < .1) or did not show the predefined contrast and were therefore not correlated with group patterns (**Figure 5**).

Participants were characterized as fitting the responder pattern if they showed a *meaningful decrease in coarse scale MSE from baseline to week 1* in their first LV, and characterized as fitting the non-responder pattern if they demonstrated *no meaningful change in coarse scale MSE from baseline to week 1, and any change in coarse scale MSE from week 1 to week 12* in their second LV. Again, two raters examined the significant patterns found in each individual analysis and independently decided whether they conformed to these definitions. Inter-rater reliability yielded ***κ***=0.91 for rating whether patients fit the responder pattern, and ***κ***=0.96 for ratings on whether patients fit the non-responder pattern. Based on consensus, 46.5% of patients exclusively showed the pattern of their own group, 25.6% fit both patterns, 14.0% exclusively showed the opposite pattern, and 14.0% showed neither. Patients in this last group, including 4 responders and 2 non-responders, showed an early *increase* in coarse scale complexity instead. The characteristics of patients rated as conforming to their own group pattern exclusively versus those who did not are presented in **Table 3**. Again, the most notable difference was that responders exclusively showing the responder pattern exhibited higher correlations between their individual and group MSE patterns compared to responders assigned to the other categories; this comparison did not survive Holm’s sequential Bonferroni test to correct for multiple comparisons (p = .006).

**Table 3.** Demographic and Clinical Characteristics (Means ± Standard Error) of Antidepressant Treatment Responders and Non-responders Divided Based on which Individuals Matched the Group MSE Patterns

|  | **Responders**<br>**(N = 25)** |  | **Non-responders**<br>**(N = 18)** |  |
| --- | --- | --- | --- | --- |
|  | *Responder pattern*<br>*only (N = 12)* | *Other categories*<br>*(N = 13)* | *Non-responder pattern*<br>*only (N = 8)* | *Other categories*<br>*(N = 10)* |
| *Sex (F/M)* | 5/7 | 9/4 | 3/5 | 6/4 |
| *Age* | 30.6 ± 3.2<br>(range: 21-46) | 38.5 ± 2.4<br>(range: 19-57) | 46.5 ± 4.0<br>(range: 33-63) | 43.4 ± 3.9<br>(range: 20-57) |
| *Education (years)* | 15.8 ± 0.5 | 15.0 ± 0.8 | 16.0 ± 0.9 | 16.6 ± 0.9 |
| *Ethnicity* | 11 Caucasian; 1 Asian | 11 Caucasian; 1 Asian; 1 South Asian | 7 Caucasian; 1 African | 10 Caucasian |
| *Comorbid anxiety (Yes/No)* | 1/11 | 2/13 | 1/7 | 1/9 |
| *Treatment regimen*<br>*(ESC+BUP/BUP+placebo/*<br>*ESC+placebo)* | 7/2/3 | 5/4/4 | 0/4/4 | 5/2/3 |
| *Baseline MADRS score* | 28.8 ± 1.4 | 29.9 ± 1.3 | 32.0 ± 1.5 | 32.3 ± 1.7 |
| *MADRS score at 1 week* | 20.3 ± 2.5 | 25.8 ± 1.9 | 27.5 ± 2.4 | 28.2 ± 2.9 |
| *MADRS score at 12 weeks* | 4.4 ± 1.1 | 7.5 ± 1.5 | 24.5 ± 2.4 | 25.2 ± 2.9 |
| *Correlation with own group*<br>*MSE pattern (>.4/<.4)* | 9/3* | 3/10 | 6/2 | 4/6 |
| *Correlation with own group*<br>*MSE pattern* | 0.59 ± 0.10* | 0.10 ± 0.13 | 0.51 ± 0.23 | 0.32 ± 0.12 |
| *Correlation with own group*<br>*WPLI pattern* | 0.44 ± 0.15 | 0.26 ± 0.16 | 0.25 ± 0.12 | 0.48 ± 0.07 |

Group differences were examined using independent samples t-tests and Fisher's exact test in Excel. \*
Group differences with $p < .05$ that did not survive the Holm's sequential Bonferroni correction for multiple comparisons. F = female, M = male, ESC = escitalopram, BUP = bupropion, MADRS = Montgomery-Åsberg Depression Rating Scale

## Discussion

The current study examined the extent of individual deviation from group results in EEG characteristics (connectivity/complexity) related to antidepressant treatment response in a typical dataset. Although our group analyses differentiated pharmacotherapy responders/non-responders overall, single-subject analyses revealed that differentiating group features only existed unambiguously in up to 61% of individuals. This suggests that, although group analyses are able to detect neural characteristics of treatment success that apply to certain patients at an individual level, a substantial proportion of individuals is poorly represented by such analyses. Critically, these findings indicate that group analyses may be insufficient for determining reliable EEG characteristics of treatment response in individual patients. A more nuanced approach, focused on individual patients, will likely be needed when applying brain-based markers of response in clinical practice.

For both EEG connectivity and complexity, our group findings were in line with some previous findings, but not others. Namely, responders showed a decrease in alpha connectivity over treatment, most notably in left fronto-temporal and right occipito-parietal electrode pairs, similar to Iseger et al. and Lee et al. [6, 7]. Contrarily, increased alpha connectivity has also been observed in response to antidepressant pharmacotherapy [9]. In line with the work of Olbrich et al. [9], we observed beta connectivity increases with successful treatment in left central, parietal and frontal areas, while others did not [6]. In line with yet others’ work [17], we found that EEG complexity at lower temporal resolution (20-40ms) decreased with treatment, and this was prominent only in responders. This differs from findings highlighting a decrease in complexity at high temporal resolutions instead [16, 17].

Together, these variable group-level findings and the substantial individual variation found in our single-subject analyses provide a possible explanation as to why reliable EEG characteristics associated with antidepressant treatment response have not yet emerged. Depending on the EEG characteristic and how it was MSE identified, 39-61% of our sample did not unambiguously show the same outcomes as their group, which is consistent with the idea that multiple response patterns to antidepressant treatment exist [50]. Taking alpha connectivity as an example, recent work shows that alpha connectivity profiles may differentially predict response to placebo versus antidepressant pharmacotherapy (setraline; [51]), highlighting the possibility that different alpha connectivity patterns may distinguish responders to different types of interventions. The decrease in alpha connectivity in treatment responders using group analyses in this and previous studies might thus only represent one of several response profiles that exist in patients with MDD.

The responder group pattern we report here involved a decrease in alpha connectivity and coarse scale complexity, and an increase in beta connectivity. Although increased alpha connectivity in patients with MDD compared to healthy controls has been interpreted in varying ways [7, 52], the decrease in alpha connectivity with successful treatment reported here supports the notion that this feature plays a key role in MDD pathology and its treatment. The importance of this frequency band is further highlighted by frequent findings of altered alpha power and hemispheric asymmetry (e.g. [53, 54]). Given that intra-hemispheric anterior-posterior beta connectivity has been associated with emotion regulation in healthy populations [55], increased beta over successful treatment could reflect increased top-down control over disturbed emotional processing in MDD in response to treatment [56]. Increased overall EEG complexity in MDD has been linked to recruiting more neural resources when performing an emotion processing task than healthy controls [15]. Generally, increased signal complexity has been associated with a greater number of simultaneously activated systems [57]. As MDD has been associated with reduced ability to suppress default mode network (DMN) activation and greater interconnectedness between affective and other information processing systems [58, 59], the decrease in complexity over successful treatment found here might indicate decreased dominance and interference by emotional processing circuits.

While our findings are in keeping with multiple response patterns existing within the population of patients with MDD, they do not provide proof of this, as we only tested whether the patterns appearing at the group level were also present in individual patients. Indeed, our approach is markedly different from other studies aiming to address applicability of neuroimaging to individual patients. For example, clustering approaches aim to divide patients into functionally relevant MDD subcategories, and machine learning (ML) methods aim to identify features that predict individual treatment outcomes. Although encouraging work has emerged [60-62], subtyping research has had limited success so far [50], and these studies are far from perfect: Most existing research relies on study samples that are too small and homogeneous to provide reliable results [63, 64]. Additionally, findings from a recent machine learning study using a large multi-site dataset (N=1188) were not replicated, highlighting methodological challenges in analyzing high dimensional datasets using such approaches [50, 65, 66]. Thus, examining the brain characteristics associated with antidepressant treatment response likely warrants the use of multiple, complementary approaches. We propose that the use of single-subject analyses could help advance this field by determining the degree of individual variation and capturing the range of patterns present at the individual level. Future studies could take this further towards individual predictions of treatment outcomes, by building a library of individual brain patterns and associated treatment outcomes which can then be clustered and matched to new patients.

### Limitations & future directions

Despite the novelty of the presented work, certain limitations exist. Most importantly, the limited sample size led us to group patients receiving different treatment regimens (i.e. escitalopram, bupropion or both) together to maintain sufficient statistical strength for our main analyses. While our sample was fairly balanced in terms of sex, we were also unable to test for sex differences. In addition, our sample included patients with different MDD symptom subtypes (e.g. melancholic, atypical) and several patients had comorbid anxiety disorders (given the high co-occurrence of anxiety in depressed individuals, we did not see the justification for excluding these individuals). Again, this heterogeneity in the sample might contribute to the individual variation we found in the individual analyses. We did compare the characteristics of patients who were and were not rated as exclusively conforming to their own group pattern. Specifically, we explored whether different regimens, comorbidity profiles and other clinical and demographic characteristics were associated with different patterns of change in EEG connectivity and complexity (see **Table 2 & 3**). While these comparisons did not reveal obvious differences, future studies with larger samples should be conducted to explore the influence of heterogeneity of treatment and patient characteristics in more detail.

In addition, there are numerous ways to quantify similarity between individual and group patterns. The correlation procedure was objective, but also led to arbitrary limits for categorizing who did and did not match the group patterns (i.e. r ≥ .4). The independent ratings avoided setting such arbitrary limits, relying instead on human judgements of similarity according to set criteria. While inter-rater reliability was high for ratings of MSE (***κ*** = 0.91-0.96), there was less agreement for the ratings of WPLI (***κ*** = 0.57-0.76), highlighting the subjectivity of this method. The lower inter-rater reliability for WPLI was likely due to the larger number of elements included in this analysis (378^*^99 compared to 28^*^20 for MSE), or could reflect more variety in WPLI response patterns compared to MSE patterns. Finally, we applied no correction for multiple comparisons in the individual analyses, as this would have made it more difficult to find the group patterns in individuals, and therefore bias the results towards our hypotheses that there would be much individual divergence from group findings. That being said, lowering the significance threshold to α = .001 using Bonferroni did not alter our MSE findings, and only changed 16% of the individual WPLI analyses (7 patients) from being significant to being insignificant. Overall, both measures of similarity showed that there was substantial individual variation in the connectivity and complexity group patterns.

### Conclusion

Most existing work involving neural characteristics of antidepressant treatment success is based on responder/non-responder group differences. We show that substantial individual variation in EEG connectivity and complexity existed in a typical sample of patients receiving pharmacotherapy for MDD. Future research should take this individual variation into account when developing and considering the utility of EEG characteristics in informing clinical practice. The single-subject approach is one of the ways current research could be advanced towards meaningfully changing the trajectory of depressed patients obtaining treatment.

## Data Availability

All EEG data analyzed in this manuscript are available in raw and preprocessed form on OSF (https://osf.io/f6pw3/), together with the datamats that were prepared for the partial least squares analyses. The clinical and demographic data cannot be shared publicly because no ethics approval has been granted for sharing this data. Permission to access these data can be requested by contacting directly.

## Acknowledgements and funding

Patients in this study were recruited from an NIH-funded clinical trial (5R01MH077285). We acknowledge support to G.W. in the form of a Mamdani Family Foundation Graduate Scholarship and Alberta Graduate Excellence Scholarship (AGES – International), and N.J. and A.B.P. from the Natural Sciences and Engineering Council of Canada (NSERC; RGPIN-2018-06869 and RGPIN-2020-05299 respectively). We would like to thank Drs. Claude Blondeau, Pierre Tessier and Sandhaya Norris for their help with clinical assessments and diagnoses.

## References

1. Iwabuchi, S.J., et al., Localized connectivity in depression: a meta-analysis of resting state functional imaging studies. Neuroscience & Biobehavioral Reviews, 2015. 51: p. 77–86.

2. Smart, O.L., V.R. Tiruvadi, and H.S. Mayberg, Multimodal approaches to define network oscillations in depression. Biological psychiatry, 2015. 77(12): p. 1061–1070.

3. Rush, A.J., et al., Acute and longer-term outcomes in depressed outpatients requiring one or several treatment steps: a STAR^*^D report. Am J Psychiatry, 2006. 163(11): p. 1905–17.

4. Leuchter, A.F., et al., Biomarkers to predict antidepressant response. Current psychiatry reports, 2010. 12(6): p. 553–562.

5. Calhoun, V.D., et al., Prediction of Individual Differences from Neuroimaging Data. Neuroimage, 2017. 145(Pt B): p. 135–136.

6. Lee, T.-W., et al., The implication of functional connectivity strength in predicting treatment response of major depressive disorder: a resting EEG study. Psychiatry Research: Neuroimaging, 2011. 194(3): p. 372–377.

7. Iseger, T.A., et al., EEG connectivity between the subgenual anterior cingulate and prefrontal cortices in response to antidepressant medication. European Neuropsychopharmacology, 2017. 27(4): p. 301–312.

8. Khodayari-Rostamabad, A., et al., A machine learning approach using EEG data to predict response to SSRI treatment for major depressive disorder. Clinical Neurophysiology, 2013. 124(10): p. 1975–1985.

9. Olbrich, S., et al., Functional connectivity in major depression: increased phase synchronization between frontal cortical EEG-source estimates. Psychiatry Research: Neuroimaging, 2014. 222(1-2): p. 91–99.

10. Garrett, D.D., et al., Moment-to-moment brain signal variability: a next frontier in human brain mapping? Neuroscience & Biobehavioral Reviews, 2013. 37(4): p. 610–624.

11. Costa, M., A.L. Goldberger, and C.-K. Peng, Multiscale entropy analysis of biological signals. Physical review E, 2005. 71(2): p. 021906.

12. Ahmadlou, M., H. Adeli, and A. Adeli, Fractality analysis of frontal brain in major depressive disorder. International Journal of Psychophysiology, 2012. 85(2): p. 206–211.

13. Akar, S.A., et al., Nonlinear analysis of EEG in major depression with fractal dimensions. Conf Proc IEEE Eng Med Biol Soc, 2015. 2015: p. 7410–3.

14. Li, Y., et al., Abnormal EEG complexity in patients with schizophrenia and depression. Clinical Neurophysiology, 2008. 119(6): p. 1232–1241.

15. Wei, L., et al. Emotion-induced higher wavelet entropy in the EEG with depression during a cognitive task. in 2009 Annual International Conference of the IEEE Engineering in Medicine and Biology Society. 2009. IEEE.

16. Méndez, M.A., et al., Complexity analysis of spontaneous brain activity: effects of depression and antidepressant treatment. Journal of psychopharmacology, 2012. 26(5): p. 636–643.

17. Thomasson, N., et al., Nonlinear EEG changes associated with clinical improvement in depressed patients. Nonlinear Dynamics, Psychology, and Life Sciences, 2000. 4(3): p. 203–218.

18. Cukic, M., et al., Nonlinear analysis of EEG complexity in episode and remission phase of recurrent depression. Int J Methods Psychiatr Res, 2020. 29(2): p. e1816.

19. Jaworska, N., et al., Pre-treatment EEG signal variability is associated with treatment success in depression. NeuroImage: Clinical, 2018. 17: p. 368–377.

20. Goldberg, D., The heterogeneity of “major depression”. World Psychiatry, 2011. 10(3): p. 226.

21. Kessler, R., et al., Using patient self-reports to study heterogeneity of treatment effects in major depressive disorder. Epidemiology and psychiatric sciences, 2017. 26(1): p. 22–36.

22. Gordon, E.M., et al., Precision functional mapping of individual human brains. Neuron, 2017. 95(4): p. 791-807. e7.

23. First, M.B., et al., Structured clinical interview for DSM-IV-TR Axis I disorders: patient edition. 2005: Biometrics Research Department, Columbia University New York, NY.

24. Stewart, J.W., et al., Combination antidepressant therapy for major depressive disorder: speed and probability of remission. Journal of psychiatric research, 2014. 52: p. 7–14.

25. D’Avanzato, C. and M. Zimmerman, The Diagnosis and Assessment of Mood Disorders. The Oxford Handbook of Mood Disorders, 2017: p. 95.

26. Montgomery, S.A. and M. Åsberg, A new depression scale designed to be sensitive to change. The British journal of psychiatry, 1979. 134(4): p. 382–389.

27. Chatrian, G., E. Lettich, and P. Nelson, Ten percent electrode system for topographic studies of spontaneous and evoked EEG activities. American Journal of EEG technology, 1985. 25(2): p. 83–92.

28. Delorme, A. and S. Makeig, EEGLAB: an open source toolbox for analysis of single-trial EEG dynamics including independent component analysis. J Neurosci Methods, 2004. 134(1): p. 9–21.

29. Oostenveld, R., et al., FieldTrip: open source software for advanced analysis of MEG, EEG, and invasive electrophysiological data. Computational intelligence and neuroscience, 2011. 2011.

30. Stam, C.J., G. Nolte, and A. Daffertshofer, Phase lag index: assessment of functional connectivity from multi channel EEG and MEG with diminished bias from common sources. Human brain mapping, 2007. 28(11): p. 1178–1193.

31. Vinck, M., et al., An improved index of phase-synchronization for electrophysiological data in the presence of volume-conduction, noise and sample-size bias. Neuroimage, 2011. 55(4): p. 1548–1565.

32. Cohen, M.X., Analyzing neural time series data: theory and practice. 2014: MIT press.

33. McDonough, I.M. and K. Nashiro, Network complexity as a measure of information processing across resting-state networks: evidence from the Human Connectome Project. Front Hum Neurosci, 2014. 8: p. 409.

34. Costa, M., A.L. Goldberger, and C.K. Peng, Multiscale entropy analysis of complex physiologic time series. Phys Rev Lett, 2002. 89(6): p. 068102.

35. Heisz, J.J., et al., A trade-off between local and distributed information processing associated with remote episodic versus semantic memory. Journal of cognitive neuroscience, 2014. 26(1): p. 41–53.

36. Richman, J.S. and J.R. Moorman, Physiological time-series analysis using approximate entropy and sample entropy. American Journal of Physiology-Heart and Circulatory Physiology, 2000. 278(6): p. H2039–H2049.

37. McIntosh, A.R., et al., The development of a noisy brain. Archives italiennes de biologie, 2010. 148(3): p. 323–337.

38. McIntosh, A., et al., Spatiotemporal dependency of age-related changes in brain signal variability. Cerebral Cortex, 2014. 24(7): p. 1806–1817.

39. Smit, D.J., et al., The brain matures with stronger functional connectivity and decreased randomness of its network. PLoS one, 2012. 7(5).

40. Vysata, O., et al., Age-related changes in EEG coherence. Neurologia i neurochirurgia polska, 2014. 48(1): p. 35–38.

41. Szostakiwskyj, J.M., et al., The modulation of EEG variability between internally-and externally-driven cognitive states varies with maturation and task performance. PloS one, 2017. 12(7).

42. Wang, H., et al., The relation between Scrabble expertise and brain aging as measured with EEG brain signal variability. Neurobiology of aging, 2018. 69: p. 249–260.

43. Allen, E.A., et al., EEG Signatures of Dynamic Functional Network Connectivity States. Brain Topogr, 2018. 31(1): p. 101–116.

44. Barry, R.J., et al., EEG differences between eyes-closed and eyes-open resting conditions. Clinical Neurophysiology, 2007. 118(12): p. 2765–2773.

45. Ibáñez-Molina, A.J., et al., Multiscale Lempel–Ziv complexity for EEG measures. Clinical Neurophysiology, 2015. 126(3): p. 541–548.

46. Tan, B., et al., The difference of brain functional connectivity between eyes-closed and eyes-open using graph theoretical analysis. Computational and mathematical methods in medicine, 2013. 2013.

47. McIntosh, A., et al., Spatial pattern analysis of functional brain images using partial least squares. Neuroimage, 1996. 3(3): p. 143–157.

48. McIntosh, A.R. and N.J. Lobaugh, Partial least squares analysis of neuroimaging data: applications and advances. Neuroimage, 2004. 23: p. S250–S263.

49. Akoglu, H., User’s guide to correlation coefficients. Turkish journal of emergency medicine, 2018. 18(3): p. 91–93.

50. Beijers, L., et al., Data-driven biological subtypes of depression: systematic review of biological approaches to depression subtyping. Molecular psychiatry, 2019. 24(6): p. 888–900.

51. Rolle, C.E., et al., Cortical connectivity moderators of antidepressant vs placebo treatment response in major depressive disorder: secondary analysis of a randomized clinical trial. JAMA psychiatry, 2020.

52. Leuchter, A.F., et al., Resting-state quantitative electroencephalography reveals increased neurophysiologic connectivity in depression. PLoS One, 2012.

53. Baskaran, A., R. Milev, and R.S. McIntyre, The neurobiology of the EEG biomarker as a predictor of treatment response in depression. Neuropharmacology, 2012. 63(4): p. 507–13.

54. Mumtaz, W., et al., Review on EEG and ERP predictive biomarkers for major depressive disorder. Biomedical Signal Processing and Control, 2015. 22: p. 85–98.

55. Miskovic, V. and L.A. Schmidt, Cross-regional cortical synchronization during affective image viewing. Brain research, 2010. 1362: p. 102–111.

56. Rive, M.M., et al., Neural correlates of dysfunctional emotion regulation in major depressive disorder. A systematic review of neuroimaging studies. Neuroscience & Biobehavioral Reviews, 2013. 37(10): p. 2529–2553.

57. Tononi, G. and G.M. Edelman, Consciousness and complexity. science, 1998. 282(5395): p. 1846–1851.

58. Anticevic, A., et al., The role of default network deactivation in cognition and disease. Trends in cognitive sciences, 2012. 16(12): p. 584–592.

59. Epstein, J., et al., Failure to segregate emotional processing from cognitive and sensorimotor processing in major depression. Psychiatry Research: Neuroimaging, 2011. 193(3): p. 144–150.

60. Iosifescu, D.V., R.J. Neborsky, and R.J. Valuck, The use of the Psychiatric Electroencephalography Evaluation Registry (PEER) to personalize pharmacotherapy. Neuropsychiatric disease and treatment, 2016. 12: p. 2131.

61. Iniesta, R., D. Stahl, and P. McGuffin, Machine learning, statistical learning and the future of biological research in psychiatry. Psychological medicine, 2016. 46(12): p. 2455–2465.

62. Wu, W., et al., An electroencephalographic signature predicts antidepressant response in major depression. Nature Biotechnology, 2020: p. 1–9.

63. Patel, M.J., A. Khalaf, and H.J. Aizenstein, Studying depression using imaging and machine learning methods. NeuroImage: Clinical, 2016. 10: p. 115–123.

64. Kim, Y.-K. and K.-S. Na, Application of machine learning classification for structural brain MRI in mood disorders: Critical review from a clinical perspective. Progress in Neuro-Psychopharmacology and Biological Psychiatry, 2018. 80: p. 71–80.

65. Drysdale, A.T., et al., Resting-state connectivity biomarkers define neurophysiological subtypes of depression. Nature medicine, 2017. 23(1): p. 28.

66. Dinga, R., et al., Evaluating the evidence for biotypes of depression: attempted replication of Drysdale et. al. 2017. bioRxiv, 2018: p. 416321.

