## Supplementary material for "One size does not fit all: Single-subject analyses reveal substantial individual variation in electroencephalography (EEG) characteristics of antidepressant treatment response": Figure S1

**1. EO/EC contrast**

The task PLS analysis of WPLI including both groups (responders/non-responders), all assessment sessions (baseline, weeks 1 & 12) and both testing states (EC, EO) identified two significant LVs (LV1: *p*<.001, PVE=42.47%; LV2: *p*=.024, PVE=15.44%). As an interaction effect was present, two separate task PLS analyses were run for non-responder and responder groups. Each analysis found one significant LV (non-responders: *p*<.001, PVE=42.47; responders: *p*<.001, PVE=53.06%). As illustrated in **Figure S1**, for both groups, the LVs revealed a contrast between the EC and EO states that was present at all assessment sessions, most dominantly expressed in higher connectivity during the EC state at the alpha frequency (8-12Hz) in both groups. The interaction effect detected in the first analysis did not survive statistical testing in the separate group analyses.

The same analysis but for MSE identified one significant LV (*p*<.001, PVE=93.34%). This LV again revealed a contrast between the EC and EO states, with higher coarse scale and lower fine scale variability present during the EO compared to the EC state in both groups (**Figure S2**).

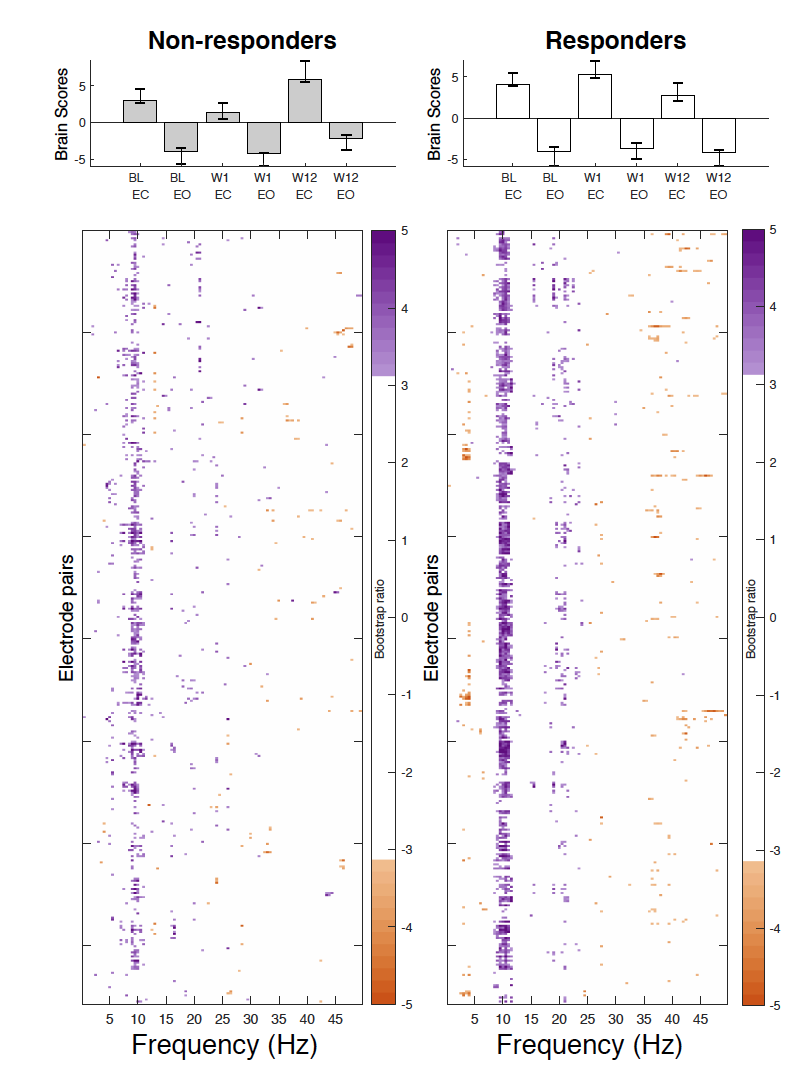
**Figure S1.** Results from the task partial-least squares (PLS) analyses examining differences in connectivity as measured by weighted phase lag index (WPLI) between eyes open (EO) and eyes closed (EC) conditions and assessment sessions (Baseline – BL, Week 1 – W1, Week 12 – W12) for non-responders (left) and responders (right). Bar graphs (A) depict the contrast between assessment sessions and EO/EC conditions within groups, that was significantly expressed across each data set as determined by permutation testing. The statistical image plots (B) present the bootstrap ratio maps over all electrode pairs (rows) and frequencies (columns). The colored values display where the contrast represented by the bar graphs was most consistent across participants as determined by bootstrapping. Positive values (purple) indicate increased WPLI during EC, while negative values (orange) indicate increased WPLI during EO.

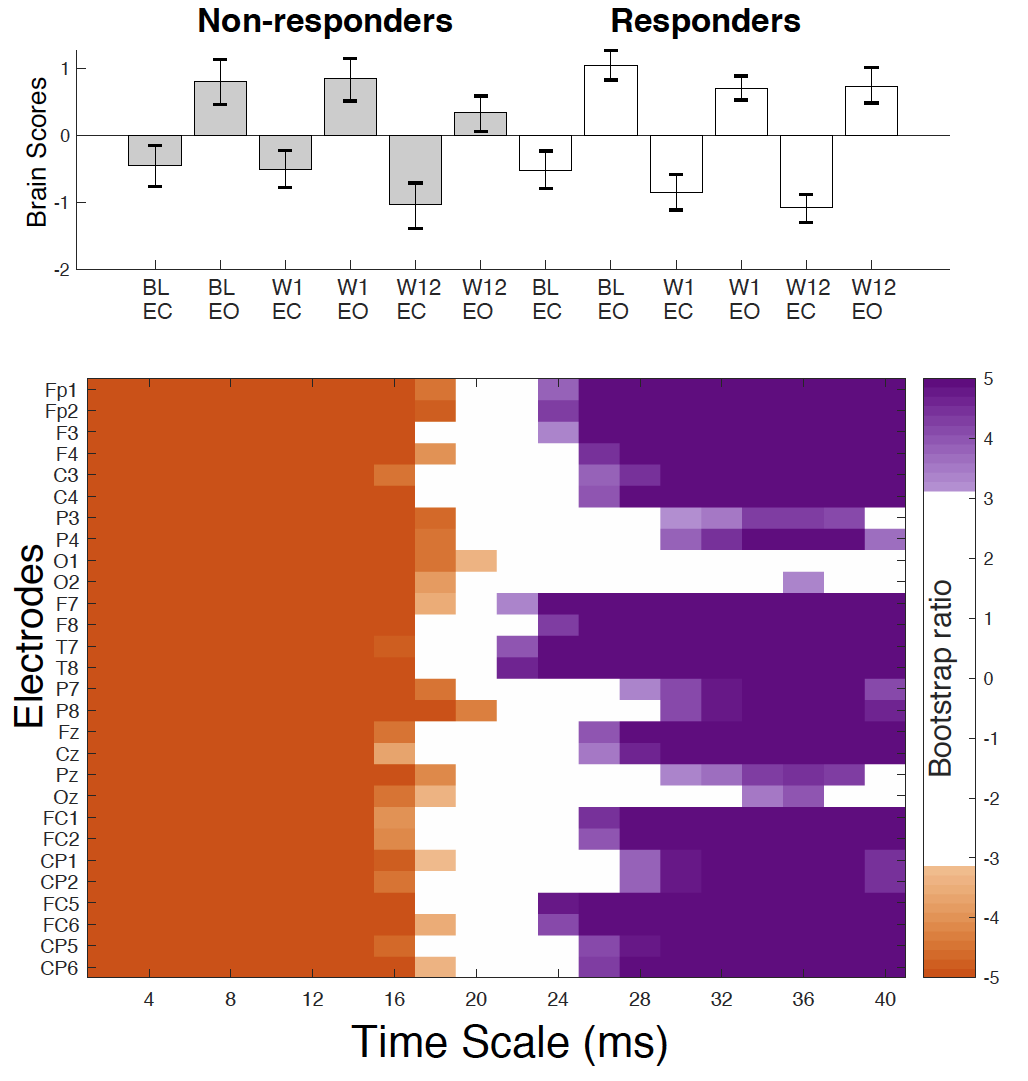

**Figure S2.** Results from the task partial-least squares (PLS) analyses examining differences in complexity as measured by multiscale entropy (MSE) between eyes open (EO) and eyes closed (EC) conditions and assessment sessions (Baseline – BL, Week 1 – W1, Week 12 – W12) for non-responders (left) and responders (right). Bar graphs depict the contrast between assessment sessions and EO/EC conditions that was significantly expressed across each data set as determined by permutation testing. The statistical image plots (B) present the bootstrap ratio maps over all electrodes (rows) and timescales (columns). The colored values display where the contrast represented by the bar graphs was most consistent across participants as determined by bootstrapping. Positive values (purple) indicate increased MSE during EO, while negative values (orange) indicate increased MSE during EC.

**2. EO results for WPLI, EC results for MSE**

*Connectivity of the EO data (task PLS) - WPLI*

The task PLS analysis of WPLI for the EO data, including both groups and all assessment sessions, identified one significant LV (*p*=.042, PVE=28.82%). As this analysis revealed an interaction effect, two additional task PLS analyses were run, one for responders and one for non-responders. Neither of these found a significant LV, although one LV showed a trend for responders (*p*= .058, PVE = 54.50%). This LV was expressed as a change from week 1 to 12 in connectivity scattered across electrode pairs and
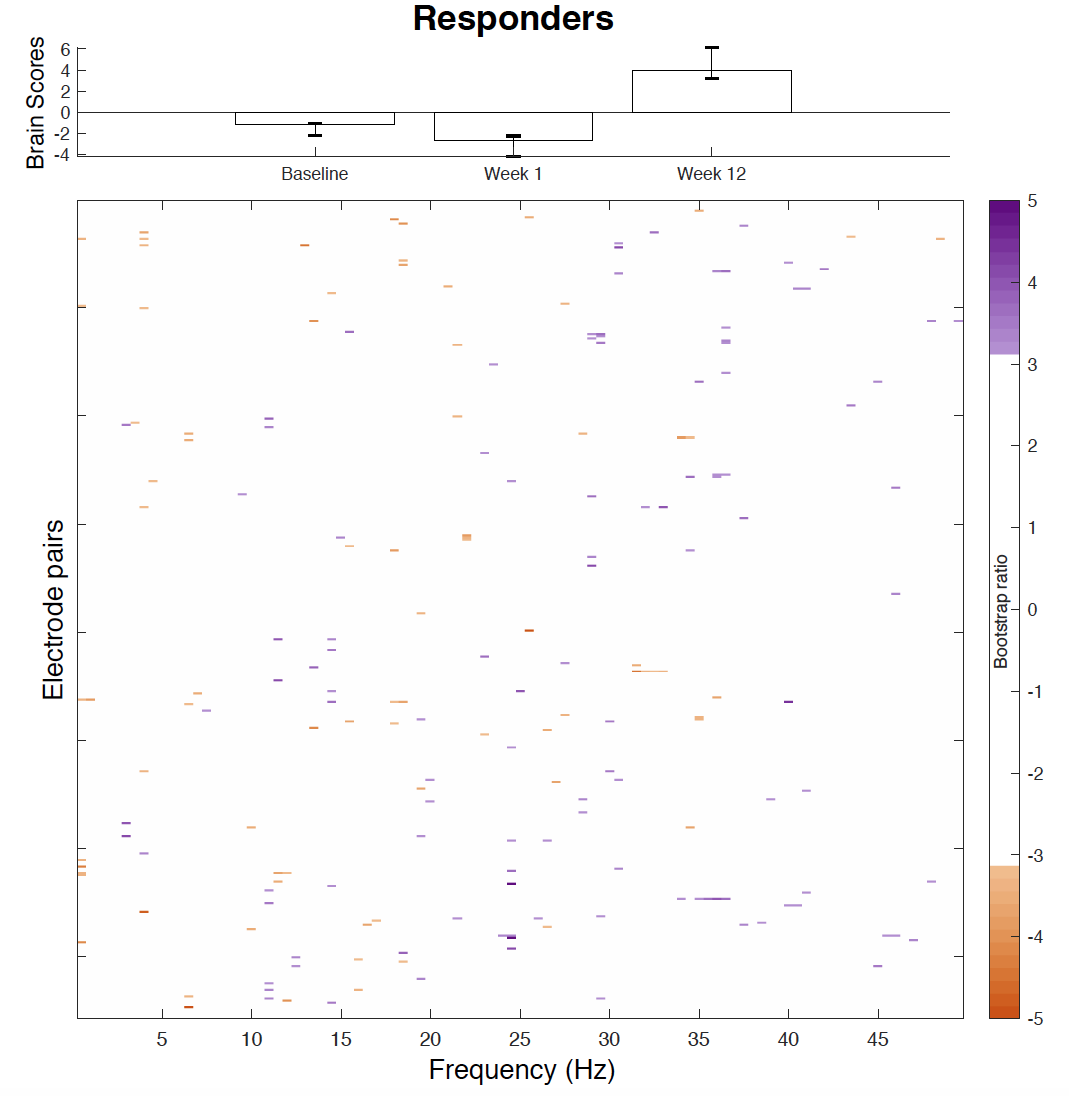
frequencies (**Figure S3**).

**Figure S3.** Results from the task partial-least squares (PLS) analyses examining change in connectivity as measured by weighted phase lag index (WPLI) over the course of antidepressant medication treatment in responders (right) during the eyes open condition. Bar graphs (A) depict the contrast between assessment sessions, that was significantly expressed across each data set as determined by permutation testing. The statistical image plots (B) present the bootstrap ratio maps over all electrode pairs (rows) and frequencies (columns). The colored values display where the contrast represented by the bar graphs was most consistent across participants as determined by bootstrapping. Positive values (purple) indicate increased WPLI from baseline/week 1 to week 12 of treatment, while negative values (orange) indicate decreased WPLI from baseline/week 1 to week 12.

*Complexity of the EC data (task PLS) - MSE*

The task PLS analysis investigating the changes in MSE across the three assessment sessions in both groups identified one significant LV (*p*=.004, PVE=66.96%). An interaction effect similar to that found with the EO data was observed, and therefore two additional task PLS analyses were performed for the responder and non-responder groups separately. For non-responders, one significant LV was identified (*p*=.042, PVE=85.58%), which revealed no change from baseline to week 1, then an increase in fine scale MSE from week 1 to 12 (left side of **Figure S4**). The LV that explained most variance for responders did not reach significance (*p*=.104, PVE=76.79%), but is illustrated to show the similarity of the effect to that found in the EO data. This LV showed a decrease in coarse scale MSE from baseline to week 1 at several electrodes, as well as an increase in fine scale MSE at two frontal electrodes in responders (right side of
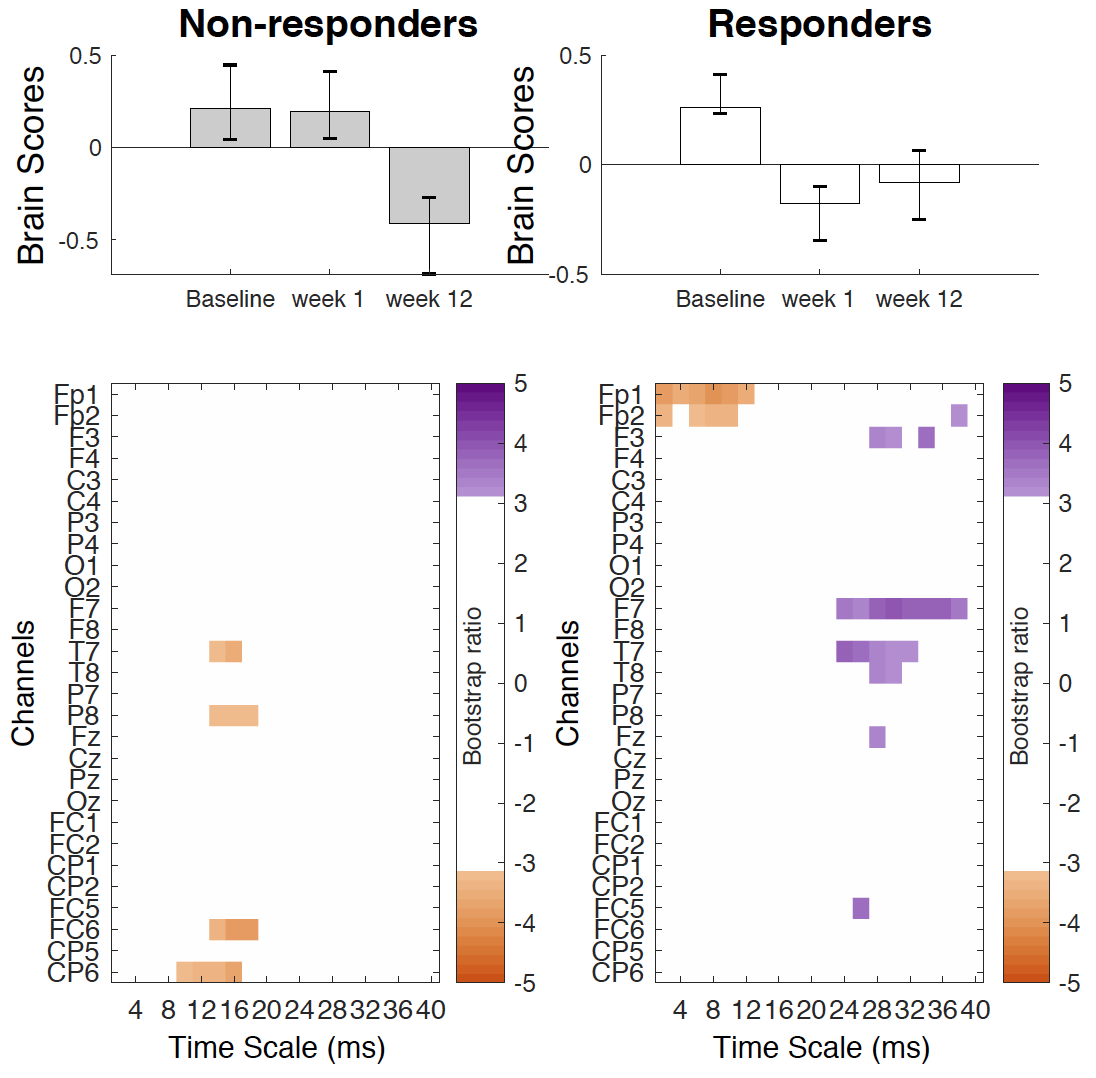
**Figure S4**).

**Figure S4.** Results from the task partial-least squares (PLS) analyses examining change in complexity as measured by multiscale entropy (MSE) over the course of antidepressant medication treatment in non-responders (left) and responders (right) during the eyes closed condition. Bar graphs (A) depict the contrast between assessment sessions within groups, that was significantly expressed across each data set as determined by permutation testing. The statistical image plots (B) present bootstrap ratio maps over all electrodes (rows) and time scales (columns). The colored values display where the contrast represented by the bar graphs was most consistent across participants as determined by bootstrapping. Positive values (purple) indicate decreased MSE, while negative values (orange) indicate increased MSE at week 12 compared to baseline and week 1 in non-responders, and at week 1 compared to baseline in responders.

**3. Group PLS analyses of single-epoch WPLI data**

As for the across-epoch WPLI data, a task PLS analysis of the averaged single-epoch WPLI data was performed with both groups and all assessments sessions (EC data only). This analysis identified one significant LV (*p*=.028, PVE=52.09%). As this LV revealed an interaction effect, two additional task PLS analyses were run for responders and non-responders separately. Of these analyses, only that for responders trended towards significance (*p*=.076, PVE=74.71%). To illustrate the similarity of these effects with the across-epoch WPLI data, the LVs explaining the most variance for each group are visually presented in **Figure S5**. As for the across-epoch WPLI data, both groups showed a change from week 1 to 12 exclusively, and this was expressed as decreased connectivity in alpha (8-13 Hz), and increased connectivity in beta (14-30 Hz) in responders, while the direction of change was opposite for non-responders. These findings were thus similar to those found in the across-epoch WPLI data, except that the results spread over multiple frequencies (e.g. from 8-13 Hz instead of dominantly at 10 Hz), which is unsurprising, considering the reduced spectral resolution associated with sliding window approaches (Cohen, 2014).

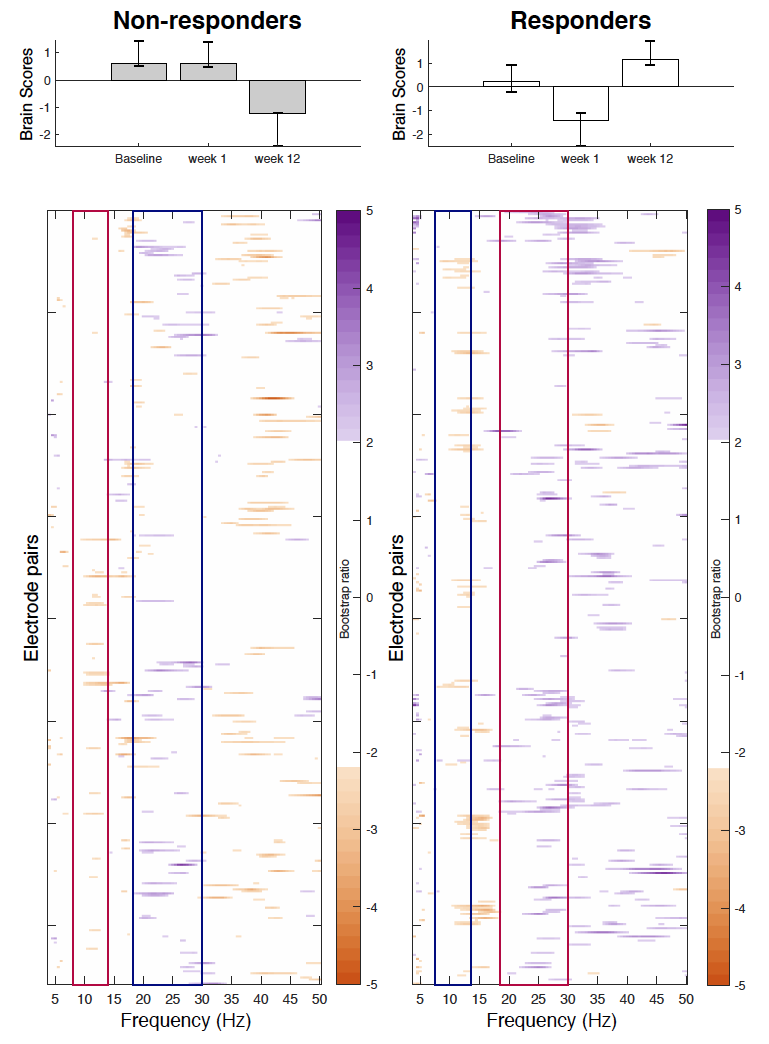
**Figure S5.** Results from the task partial-least squares (PLS) analyses examining change in connectivity as measured by averaged single-epoch weighted phase lag index (WPLI) over the course of antidepressant medication treatment in non-responders (left) and responders (right). Bar graphs (A) depict the contrast between assessment sessions within groups, as that was significantly expressed across each data set determined by permutation testing. The statistical image plots (B) present the bootstrap ratio maps over all electrode pairs (rows) and frequencies (columns). The colored values display where the contrast represented by the bar graphs was most consistent across participants as determined by bootstrapping. Positive values (purple) indicate increased WPLI in responders, and decreased WPLI in non-responders from 1 to 12 weeks of treatment, while negative values (orange) indicate decreased WPLI in responders and increased WPLI in non-responders from weeks 1 to 12. To aid interpretability, the frequency bands showing the most prominent increases in WPLI in the main WPLI analysis are highlighted by red boxes, while decreases in WPLI are outlined by blue boxes. As highlighted by these boxes, non-responders showed an increase in alpha and a decrease in beta connectivity from week 1 to week 12 of treatment, while responders showed the opposite pattern.
